# Comparison of electrophysiological left bundle branch pacing characteristics in different bilateral electrode pacing vector configurations

**DOI:** 10.1101/2023.12.05.23299529

**Authors:** Hao Wu, Longfu Jiang, Jiabo Shen, Lu Zhang

**Author notes:** **Corresponding author:** Longfu Jiang, MD, Professor, Department of Cardiovascular Medicine, Ningbo NO.2 Hospital, Ningbo City, Zhejiang province, China., Cardiovascular Disease Clinical Medical Research Center of Ningbo, Zhejiang, China. Address: NO.41, Northwest Street, Haishu District, Ningbo City, Zhejiang Province, China.

## Abstract

**Background:** Left bundle branch pacing (LBBP) in bipolar pacing produces a more balanced ventricular activation than conventional unipolar pacing but need high pacing output. The present study aimed to compare the electrophysiological characteristics of LBBP in different bilateral electrode pacing vector configurations.

**Methods:** A total of 57 patients who met the criteria for left bundle branch (LBB) capture and underwent three bilateral electrode pacing vector configuration test were enrolled. The electrocardiogram (ECG) and electrogram (EGM) parameters were evaluated and other electrophysiological characteristics were analyzed using a three-electrode configuration test.

**Results:** Seven capture modes (right ventricular septal (RVS) + left ventricular septal (LVS) + LBB, RVS + LBB, LVS + LBB, RVS + LVS, RVS, LVS, and LBB) were utilized in the study. The thresholds of full fusion mode (RVS+LVS+LB) in Bilateral Cathodes and Ring Bipolar were all lower than that in Tip Bipolar (1.2 ± 0.5 V vs. 2.7 ± 1.0 V, P < 0.001; 1.6 ± 0.6 V vs. 2.7 ± 1.0 V, P < 0.001). Full fusion mode had the shortest P-QRS (116.9 ± 12.8 ms), V6 RWPT (64.9 ± 9.7 ms), and V1 RWPT (94.5 ± 12.3 ms).

**Conclusion:** Changing the bilateral electrode pacing vector configuration to Bilateral Cathodes and Ring Bipolar can reduce the full fusion mode capture threshold compared to conventional bipolar pacing.

**What is Known?:**

1. Left bundle branch pacing in bipolar pacing can capture left bundle branch, left ventricular septal myocardial, and right ventricular septal myocardial at a higher pacing output, which was termed full fusion mode.

**What the Study Adds?:**

1. This study indicates that the full fusion mode threshold can be reduced by changing the pacing vector configuration.
2. Seven capture modes were observed in Ring Bipolar and Bilateral Cathodes during threshold testing.

GRAPHICAL ABSTRACT
The characteristics of ECG and EGM of different types of capture in three bilateral electrode pacing vector configurations and the schematic representation.

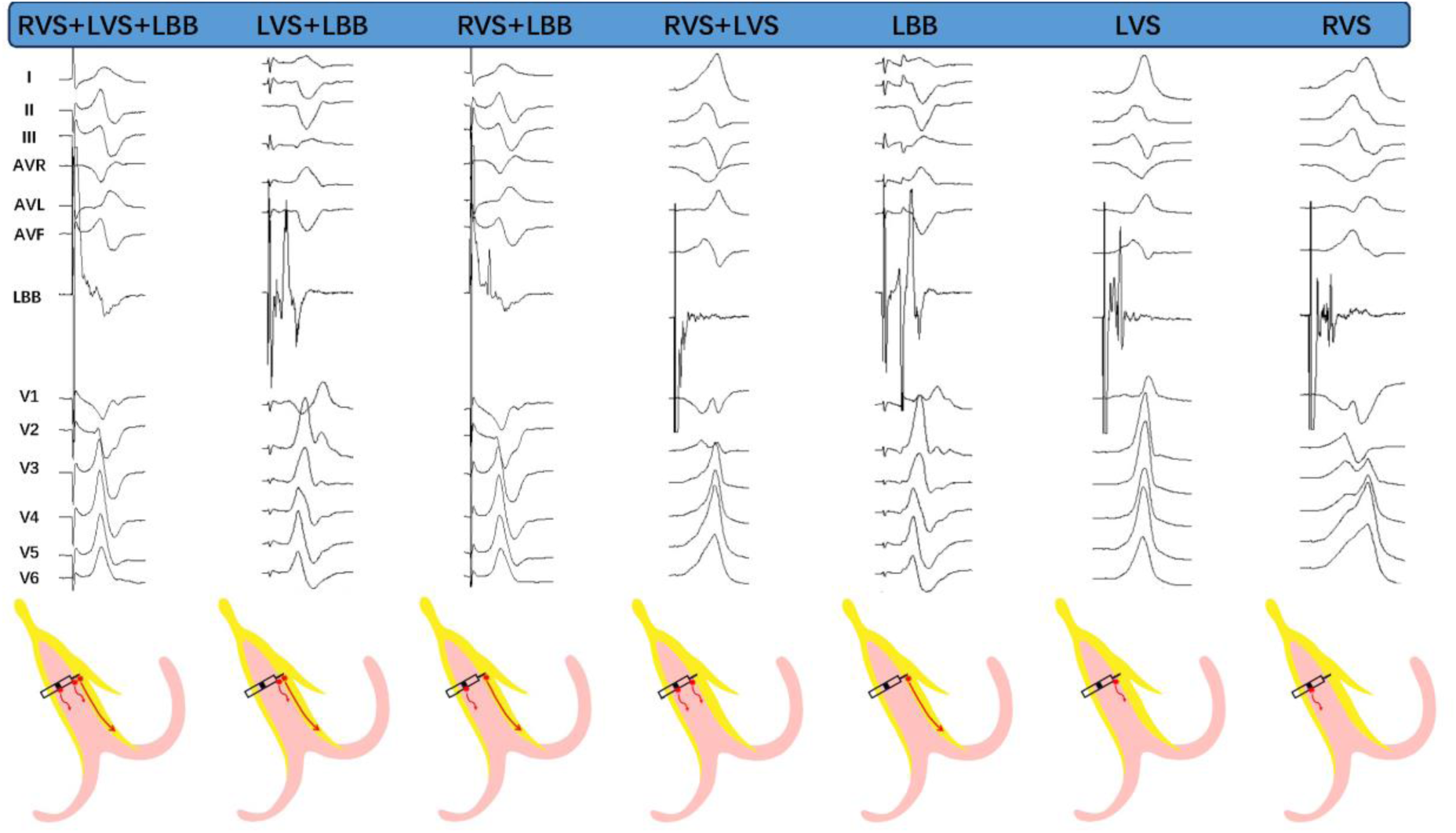

## Introduction

Conventional right ventricular (RV) pacing is a standard treatment for patients with symptomatic bradyarrhythmia, but it is associated with the risk of heart failure due to electrical and mechanical dyssynchrony.^1,2^ His bundle pacing (HBP) is a more physiological pacing modality with an improvement in exercise capacity and left ventricular (LV) ejection fraction.^3,4^ However, it also has limitations, including longer procedure times and high and sometimes unstable pacing capture threshold.^5,6^ Recently, left bundle branch pacing (LBBP) has emerged as another conduction system pacing modality with a low and stable threshold compared to HBP.^7–10^ LBBP produces a fast LV activation, but the presence of a right bundle branch block (RBBB) or even incomplete RBBB morphology remains a concern for interventricular dyssynchrony.^11^

We have discovered that LBBP in bipolar pacing can capture left bundle branch (LBB), left ventricular septal (LVS) myocardial, and right ventricular septal (RVS) myocardial at a higher pacing output when the lead tip is placed in the left septal subendocardial area and the ring electrode is in contact with the right septal endocardium,^12^ which was termed full fusion mode. This mode results in a more balanced ventricular activation than conventional unipolar pacing in LBBP.^13^ However, the full fusion mode capture threshold was high in bipolar pacing because the RVS capture threshold in bipolar pacing was much higher than that in unipolar pacing. Different RVS thresholds were likely the result of capture by either the cathode or anode. Therefore, the effect of changing the bilateral electrode pacing vectors, such as using the tip electrode as the anode or both the tip and ring electrodes as the cathodes, was investigated. The present study sought to compare the electrophysiological characteristics of LBBP in different bilateral electrode pacing vector configurations.

## Methods

### Patient population and implantation procedure

Prospective consecutive enrolled patients referred for LBBP between March 2022 and November 2022 at the Ningbo No. 2 Hospital were evaluated in the present study. The study protocol was approved by the hospital institutional review board (SL-KYSB-NBEY-2021-079-01), and all patients provided written informed consent.

A SelectSecure™ lead (model 3830, 69 cm, Medtronic Inc., Minneapolis, MN, USA) and a fixed-curve sheath (C315 HIS, Medtronic, Minneapolis, MN) were used during the lead implant procedure. The continuous LBBP pacing technique was utilized as described previously.^14–16^ Patients with direct evidence of LBB capture obtained during dynamic electrocardiogram (ECG) maneuvers as a demonstration of non-selective LBB to selective LBB capture or non-selective LBB to septal capture transition during unipolar pacing threshold testing were included in the study to ensure data accuracy.

Patients who had already undergone LBBP and three bilateral electrode pacing vector configuration testing, which was not yet a conventional procedure before March 2022, were retrospectively enrolled to increase the study sample size.

### Multiple bilateral electrode pacing vector configuration testing

Three different bilateral electrode pacing vector configurations were tested during the threshold test (Figure 1): (1) tip electrode as the cathode and ring electrode as the anode (Tip Bipolar); (2) tip electrode as the anode and ring electrode as the cathode (Ring Bipolar); and (3) both tip and ring electrodes as the cathodes (Bilateral Cathodes). There were predictable and consistent ECG and electrogram (EGM) morphology changes during the threshold test of three different bilateral electrode pacing vector configurations (Figures 2 and 3).

**FIGURE 1.**
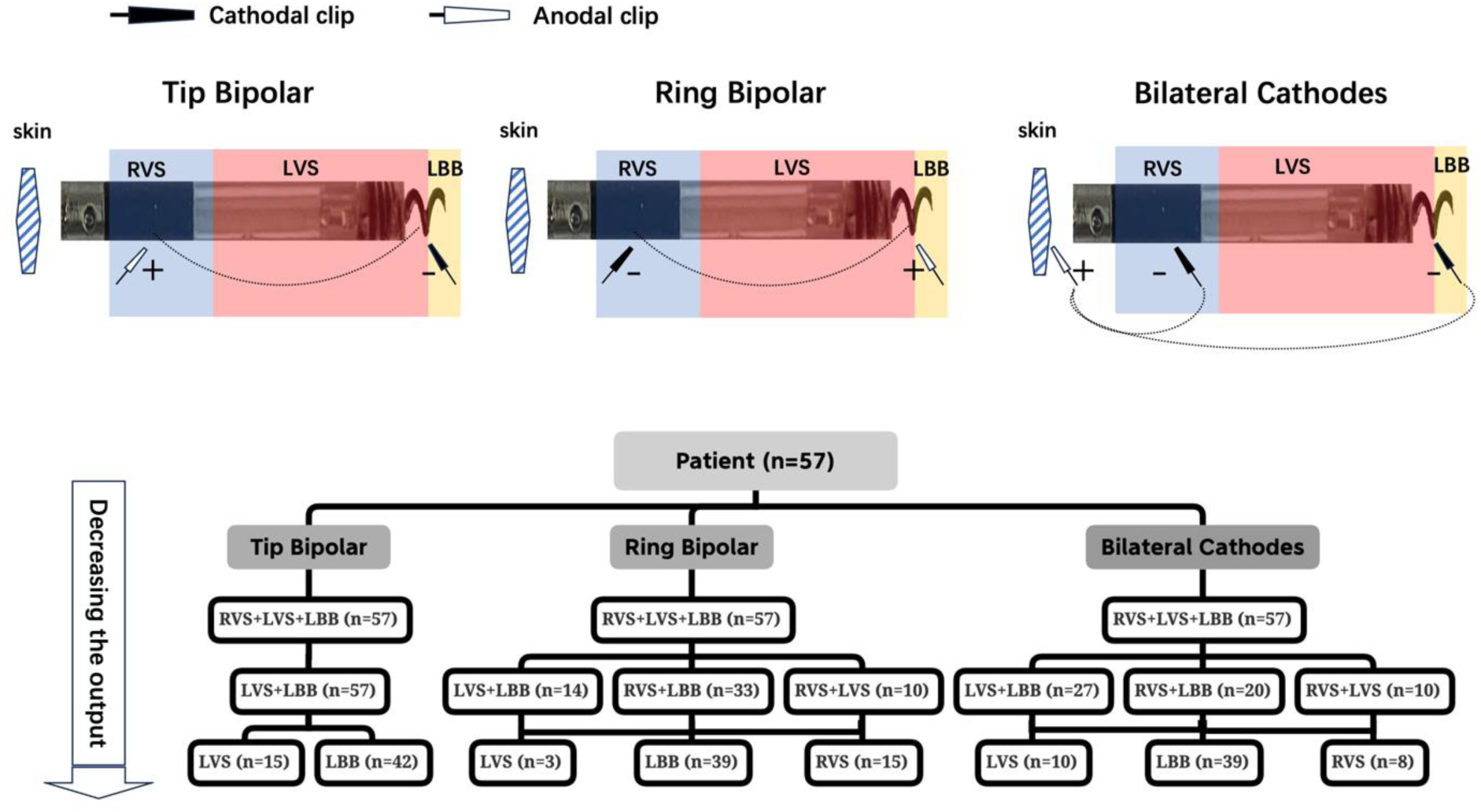
Schematic representation of three bilateral electrode pacing vector configurations and distribution of seven capture modes. “ + ” = anodal; “ - ” = cathodal. The dotted line indicates electric current flow path.

**FIGURE 2.**
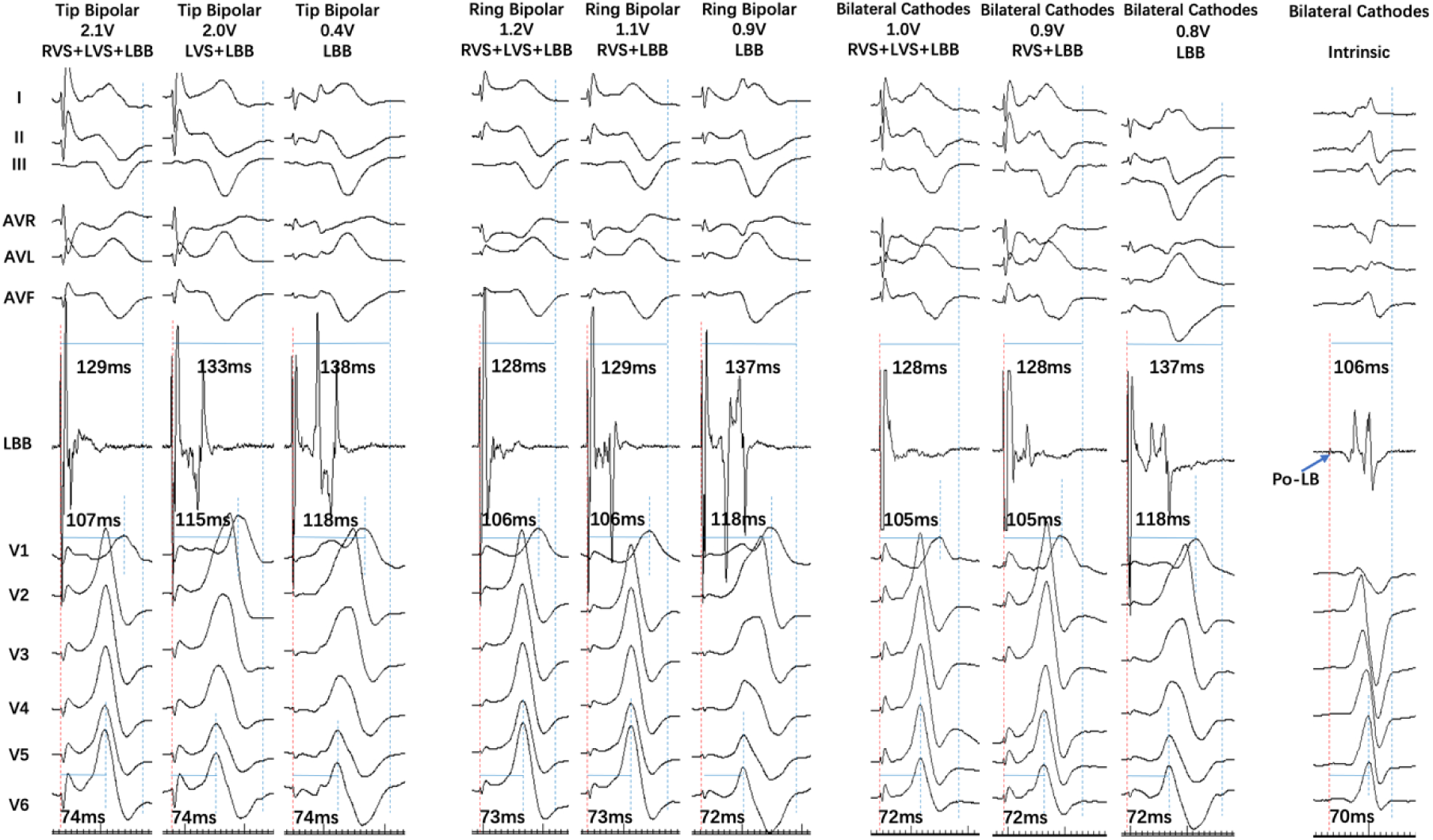
A single patient with four capture mode transitions in three bilateral electrode pacing vector configurations. In Tip Bipolar, the output decreased to 2.0 V, RVS+LVS+LBB transferred to LVS+LBB, LB lead EGM was separated, and V1 RWPT was prolonged. The output further decreased to 0.4 V, LVS+LBB transferred to LBB, V1 changed to “M” type, V1 RWPT was further prolonged, LB lead EGM was further separated, and there was an isoelectric interval between pacing artifact and local ventricular electrogram. In Ring Bipolar, the output decreased to 1.1 V, RVS+LVS+LBB transferred to RVS+LBB, ECG remained the same, and LB lead EGM was separated. The output further decreased to 0.9 V and RVS+LBB transferred to LBB. Bilateral Cathodal transition was similar to that of Ring Bipolar.

**FIGURE 3.**
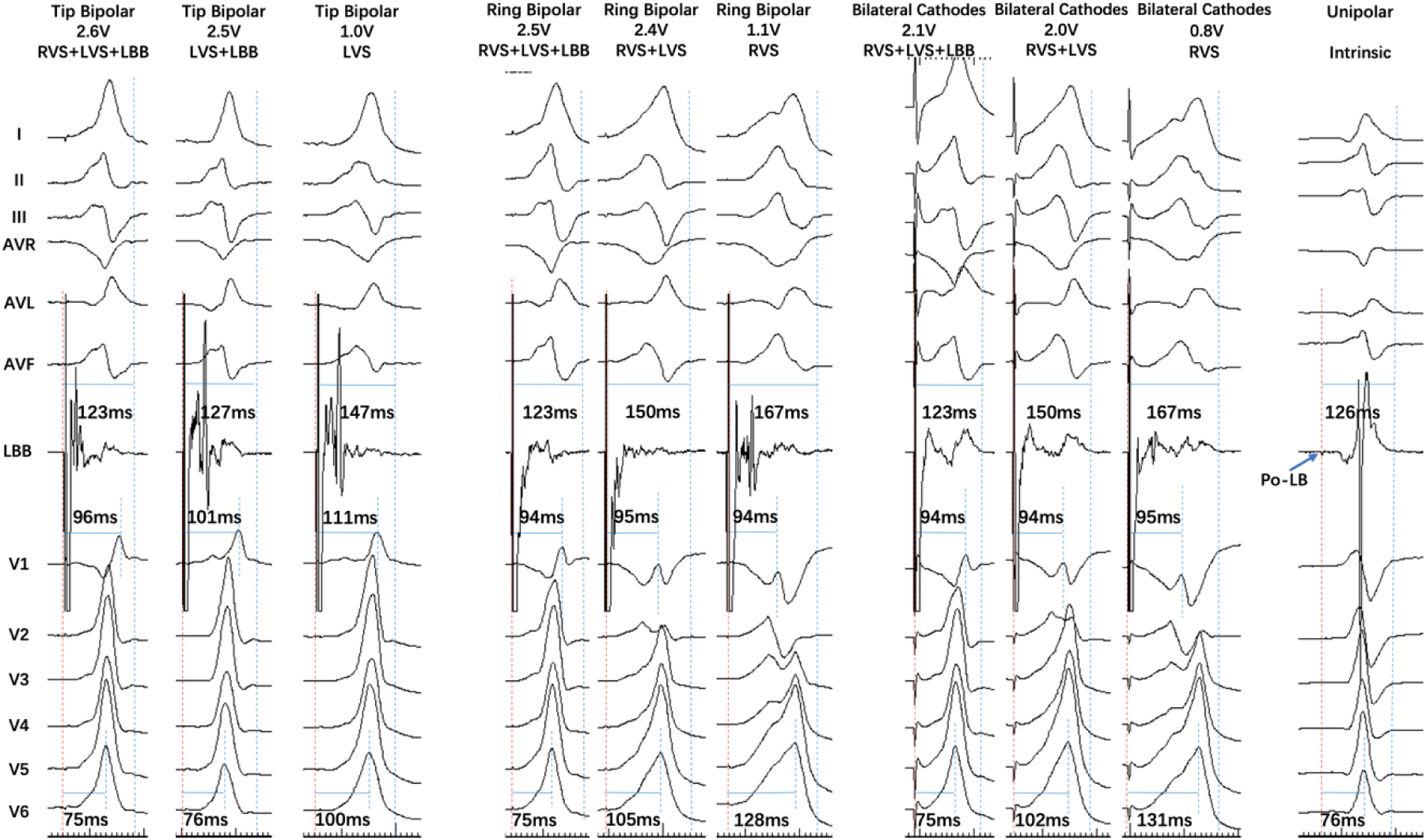
A single patient with five capture mode transitions in three bilateral electrode pacing vector configurations. In Tip Bipolar, the output decreased to 2.5 V, RVS+LVS+LBB transferred to LVS+LBB, LB lead EGM was separated, and V1 RWPT was prolonged. The output further decreased to 1.0 V, LVS+LBB transferred to LVS, LB lead EGM remained similar, and V6 RWPT was prolonged. In Ring Bipolar, the output decreased to 2.4 V, RVS+LVS+LBB transferred to RVS+LVS, LB lead EGM remained similar, and V6 RWPT was prolonged. The output further decreased to 1.1 V, RVS+LVS transferred to RVS, LB lead EGM was separated, and V6 RWPT was further prolonged. Bilateral Cathodal transition was similar to that of Ring Bipolar.

### Evaluation of ECG and EGM parameters

Our previous study distinguished four capture modes during a conventional bipolar pacing threshold test based on the changes in ECG and left bundle (LB) lead EGM.^12^ When the pacing output was reduced, LB lead EGM separation (a complex mixed wave changed to two connected individual waves) indicated one-sided ventricular myocardial loss capture. Losing the RVS capture prolonged the V1 RWPT (stimulus–peak of the R wave). Losing the LBB capture prolonged the V6 RWPT or resulted in an isoelectric interval between the pacing artifact and local ventricular EGM in selected LBB captures. Seven different capture modes can be distinguished based on this theory according to the changes in EGM and ECG during the threshold test of the three bilateral electrode pacing vector configurations (Supplementary Figure 1, Figures 2 and 3). These were: A) full fusion mode, capture RVS + LVS + LBB; B) semi fusion mode, included three capture modes, RVS + LBB, LVS + LBB, and RVS + LVS; and C) select capture mode, also included three capture modes, RVS, LVS, and LBB.

### Data collection

Baseline patient characteristics and indications for pacing were recorded in addition to echocardiographic data and baseline QRS duration. Implantation and threshold test procedures were recorded on a digital electrophysiological system (Abbott Laboratories, Chicago, IL). To ensure high precision, the measurements were performed using all 12 surface ECG leads recorded simultaneously, digital calipers, fast sweep speed (200–600 mm/s), and appropriate signal augmentation. At least three QRS complexes were measured and their values were averaged. Paced QRS (P-QRS) duration was calculated based on the pacing stimulus and the final QRS component in any of the 12 ECG leads. The presence of LBB potential and LBB potential-final QRS component duration (LB-QRS) were also recorded.

The characteristics of various changes in ECG and EGM morphology in three different bilateral electrode pacing vector configuration testing and the thresholds of seven capture modes were documented. The RWPT in surface leads V6\V1 (paced ECG morphology of V1 lead in RVS capture mode presented as QS type, so it had no V1 RWPT in RVS capture mode) and P-QRS of seven capture modes were determined.

### Statistical analysis

Continuous variables were reported as mean ± standard deviation. Categorical variables were expressed as percentages. Paired comparisons were made using the student t-test if the data were normally distributed. ANOVA was used for comparing more than two groups in repeated measurements, while LSD post hoc test was used for two-group comparisons. A p-value of <0.05 was considered statistically significant. Statistical analysis was performed using IBM SPSS Statistics for Macintosh (version 26.0, IBM Corp, Armonk, NY, USA).

## Results

### Population

A total of 72 patients underwent the LBBP procedure. Of these, 56 cases had clear dynamic ECG maneuvers during the unipolar pacing threshold test, and 53 patients underwent three different bilateral electrode pacing vector configuration testing. Four patients who have undergone three different bilateral electrode pacing vector configuration testing before March 2022 were retrospectively enrolled. A total of 57 cases were enrolled in the study. The primary clinical and procedure-related characteristics of the included patients are presented in Tables 1 and 2. The unipolar pacing threshold test showed the S-LB threshold captured by the tip electrode to be 0.5 ± 0.2 V, the LVS threshold captured by the tip electrode to be 0.6 ± 0.1 V, and the RVS threshold captured by the Ring electrode to be 0.9 ± 0.5 V.

**Table 1.** Patient characteristics (n = 57)

|  |  |
| --- | --- |
| Age (years) | 71.3 ± 10.1 |
| Male gender | 28 (49.1%) |
| Pacing indication (n) |  |
| Sick sinus syndrome | 14 (24.6%) |
| Atrioventricular block | 42 (73.7%) |
| Heart failure | 1 (1.7%) |
| Comorbidities (n) |  |
| Diabetes mellitus | 20 (35.1%) |
| Hypertension | 33 (57.9%) |
| Atrial fibrillation | 14 (24.6%) |
| Cardiomyopathy | 3 (5.3%) |
| Coronary heart disease | 8 (14.0%) |
| Heart failure | 8 (14.0%) |
| Left ventricular ejection fraction (%) | 62.1 ± 11.1 |
| Left ventricular end-diastolic dimension (mm) | 49.1 ± 5.9 |
| Native QRS type |  |
| Narrow | 40 (70.1%) |
| RBBB | 12 (21.1%) |
| LBBB | 5 (8.8%) |
| Native QRS duration (ms) | 103.8 ± 25.2 |

**Table 2.**
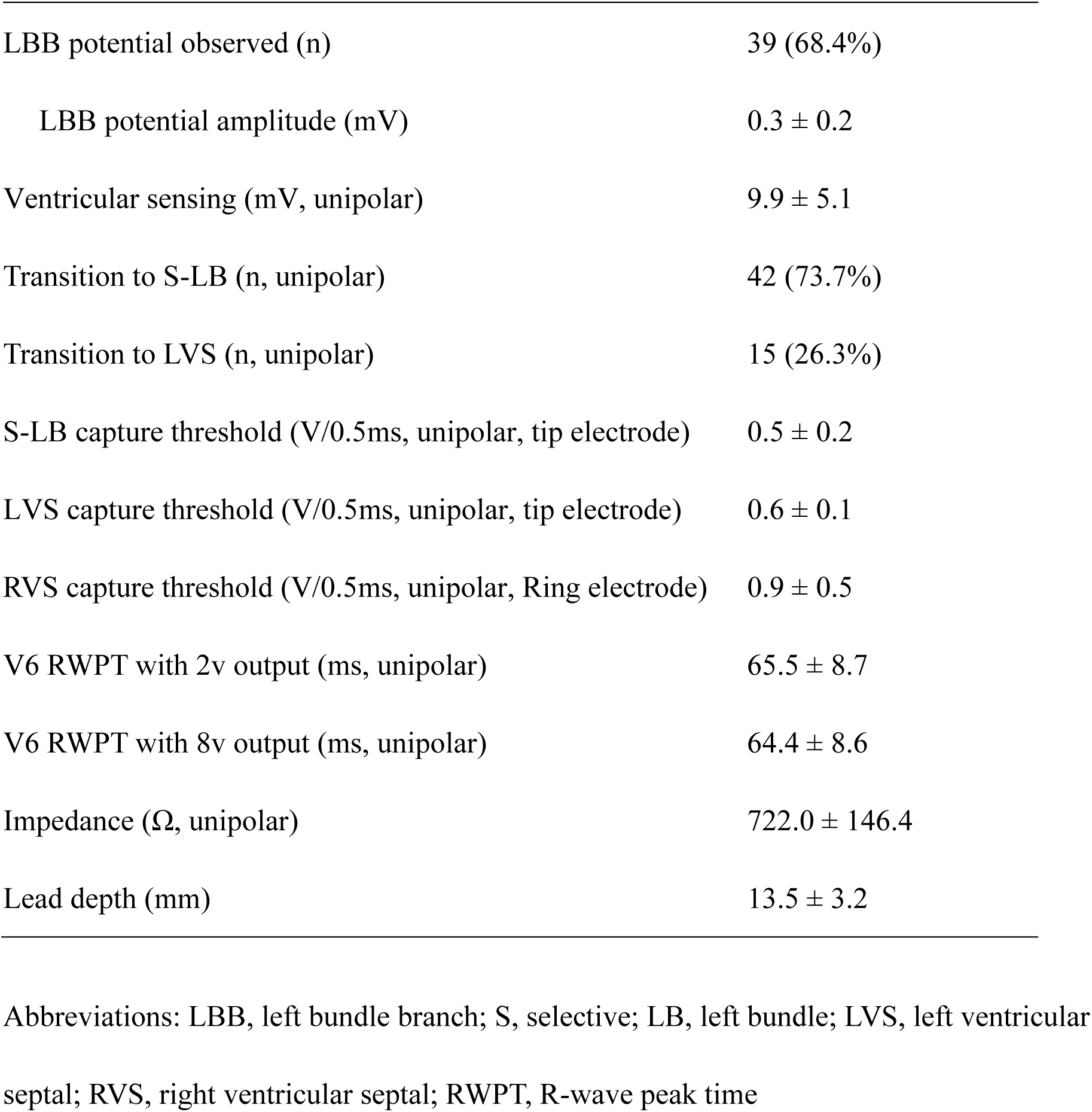
Pacing and procedure-related characteristics (n = 57)

### Transitions of seven capture modes during three bilateral electrode pacing vector configuration testing

The distribution of seven capture modes during three different bilateral electrode pacing vector configuration testing is shown in Figure 1. The transition of seven capture modes during three different bilateral electrode pacing vector configuration testing is shown in Figures 2 and 3.

The full fusion mode (RVS+LVS+LBB) was achieved in Tip Bipolar, Ring Bipolar, and Bilateral Cathodes with a higher pacing output in all 57 patients. LBB and LVS were captured using the tip side and RVS was captured using the ring side, which presented as “QS, Qr, or QR” pattern in lead V1, a complex mixed wave in LB lead EGM, short V1\V6 RWPT, and narrow P-QRS.

There are two types of transition in the Tip Bipolar configuration (conventional bipolar pacing) that were previously described.^12^ Therefore, the detailed changes in ECG and EGM are not discussed in this paper. A loss of RVS capture resulted in LBB+LVS capture mode in all 57 patients as the output decreased. A change to selected LBB capture in 42 patients and LVS capture in 15 patients occurred as the output was reduced further.

In the Ring Bipolar configuration mode, only 14 patients changed to LVS+LBB capture as the output was reduced. A total of 33 patients changed to RVS+LBB capture (EGM separated and ECG remained the same as full fusion mode), and 10 patients changed to LVS+RVS capture (V6 RWPT suddenly prolonged and EGM remained almost the same as full fusion mode). Furthermore, 39 patients changed to selected LBB capture, three patients changed to LVS capture, and 15 patients changed to RVS capture (V6 RWPT further prolonged) when the output was reduced further.

In the Bilateral Cathodes mode, a similar transition of capture modes as that in the Ring Bipolar configuration mode was observed during the threshold test. A total of 27 patients changed to LVS+LBB capture, 20 patients changed to RVS+LBB capture, and 10 patients changed to RVS+LVS capture. As the output was reduced further, 39 patients changed to LBB capture, 10 patients changed to LVS capture, and eight patients changed to RVS capture.

### Ventricular sensing, impedance, and threshold of seven capture modes in three bilateral electrode pacing vector configurations

Bilateral Cathodes mode had the lowest ventricular sensing and impedance (7.0 ± 4.3 mV; 421.4 ± 83.5 Ω; Figure 4). Full fusion mode thresholds in Bilateral Cathodes and Ring Bipolar were all lower than that in Tip Bipolar (1.2 ± 0.5 V vs. 2.7 ± 1.0 V, P < 0.001; 1.6 ± 0.6 V vs. 2.7 ± 1.0 V, P < 0.001). The threshold of LVS+LBB capture mode in Ring Bipolar mode was much higher than that in Tip Bipolar (1.1 ± 0.5 V vs. 0.7 ± 0.2 V, P < 0.001). The select LBB and LVS capture threshold in Ring Bipolar mode was much higher than that in Tip Bipolar (0.7 ± 0.2 V vs. 0.4 ± 0.2 V, P < 0.001; 1.0 ± 0.2 V vs. 0.6 ± 0.2 V, P < 0.001). There was no patient with RVS+LBB and LVS+RVS capture mode in Tip Bipolar.

**FIGURE 4.**
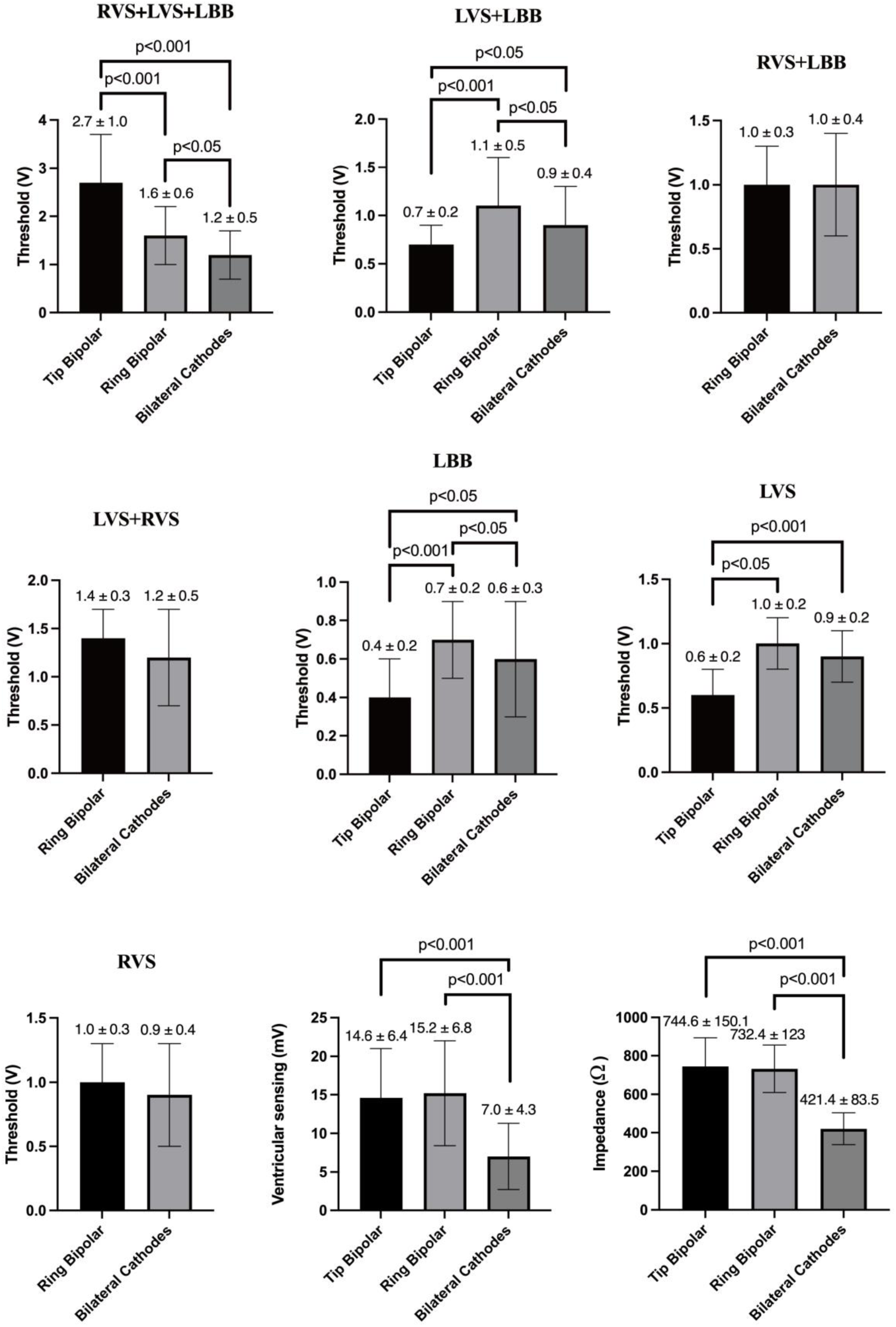
Ventricular sensing, impedance, and seven capture mode thresholds in three bilateral electrode pacing vector configurations.

### P-QRS in seven capture modes

Full fusion mode had a similar P-QRS compared to that in RVS+LBB mode (116.9 ± 12.8 ms vs. 122.1 ± 13.5 ms), but was shorter compared to those of other modes (Figure 5). The P-QRS of select RVS capture mode was the longest (143.1 ± 20.9 ms).

**FIGURE 5.**
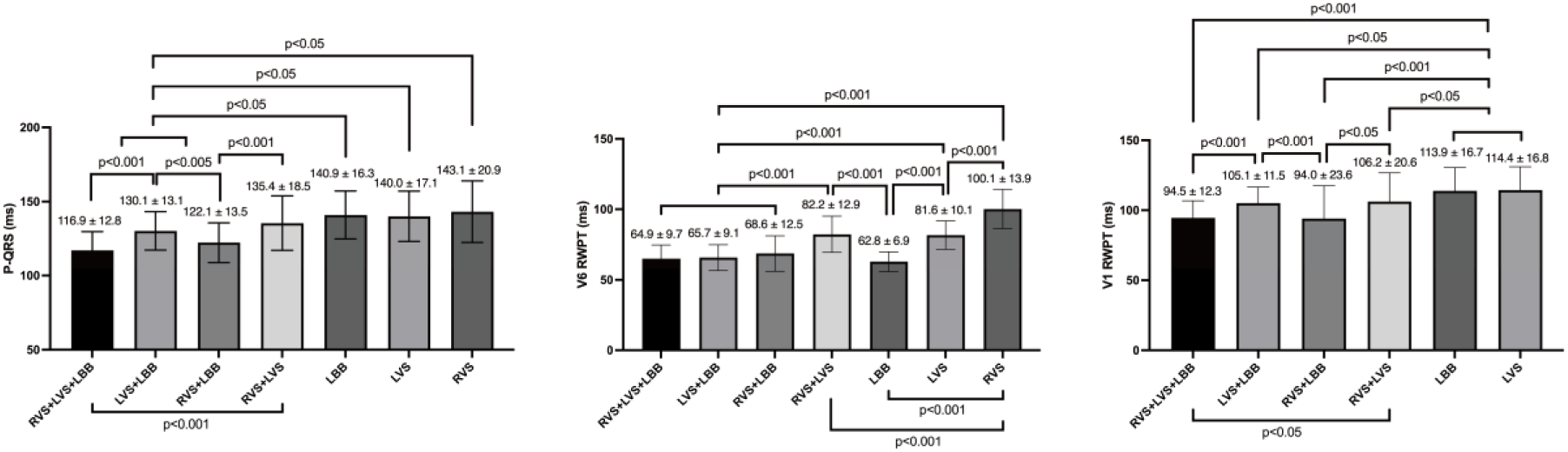
V6 RWPT, V1 RWPT, and P-QRS in seven capture modes.

### V6 RWPT in seven capture modes

Full fusion mode, LVS+LBB, RVS+LBB, and select LBB capture mode all had a short V6 RWPT and there were no significant differences among them (64.9 ± 9.7 ms vs. 65.7 ± 9.1 ms vs. 68.6 ± 12.5 ms vs. 62.8 ± 6.9 ms; Figure 5). Select RVS capture mode demonstrated a longer V6 RWPT than select LVS capture mode (100.1 ± 13.9 ms vs. 81.6 ± 10.1 ms, P < 0.001).

### V1 RWPT in seven capture modes

Full fusion mode and RVS+LBB capture mode all showed the shortest V1 RWPT (94.5 ± 12.3 ms vs. 94.0 ± 23.6 ms; Figure 5). LVS capture and select LBB capture mode had the longest V1 RWPT (114.4 ± 16.8 ms vs. 113.9 ± 16.7 ms).

### Full fusion mode in patients with native narrow and wide QRS

Full fusion mode was compared in subgroups of patients with a native narrow and wide QRS. A total of 40 patients had a native narrow QRS, and 17 patients had a native wide QRS (Figure 6). The P-QRS duration of full fusion mode was almost as short as the LB-QRS duration in intrinsic rhythm in patients with native narrow QRS (118.5 ± 10.8 ms vs. 112.8 ± 17.5 ms, P = 0.1). In patients with native wide QRS, the P-QRS duration of full fusion mode showed a statistically significant difference to the LB-QRS duration in intrinsic rhythm (120.1 ± 14.5 ms vs. 149.6 ± 19.7 ms, P < 0.05).

**FIGURE 6.**
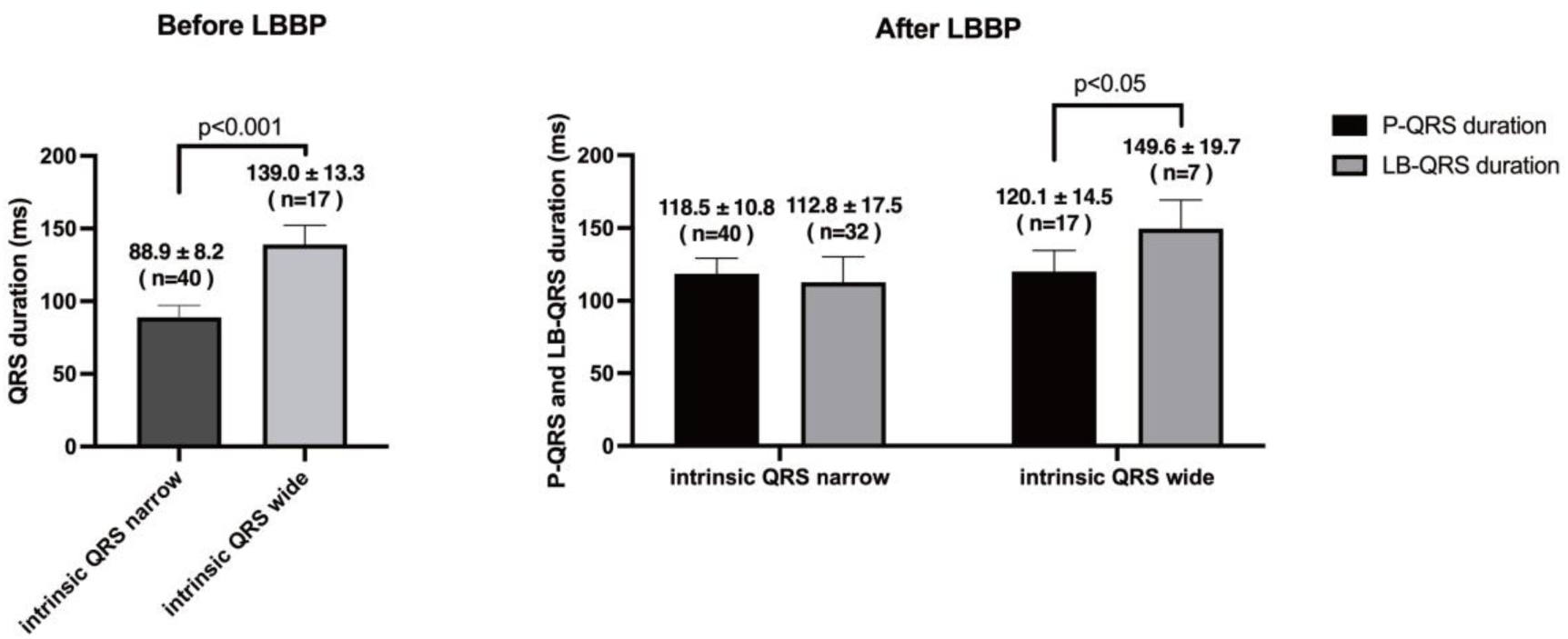
Comparison of P-QRS of full fusion mode and LB-QRS in intrinsic rhythm in patients with native narrow and wide QRS after LBBP.

## Discussion

The main findings of the present study were as follows: (1) Three bilateral electrode pacing vector configurations can all achieve full fusion mode but with different capture thresholds. Bilateral Cathodes had the lowest threshold in full fusion mode, and Tip Bipolar had the highest threshold in full fusion mode. (2) Seven capture modes were observed in Ring Bipolar and Bilateral Cathodes during threshold testing, including full fusion mode (RVS+LVSP+LBB), semi fusion mode (LVS+LBB, RVS+LBB, and RVS+LVS), and select capture mode (LBB, LVS, and RVS). (3) Full fusion mode had the shortest P-QRS, V6 RWPT, and V1 RWPT. It achieved a short P-QRS in both narrow and wide native QRS types.

### Different full fusion mode thresholds in three bilateral electrode pacing vector configurations

The present study results showed that Bilateral Cathodes had the lowest threshold in full fusion mode (1.2 ± 0.5 V), while Tip Bipolar had the highest (2.7 ± 1.0 V). This indicates that the full fusion mode threshold can be reduced by changing the pacing vector configuration. Cathodal stimulation of the cardiac septal myocardium occurs as a negative charge is applied to the myocardial–lead interface, which can direct cell depolarization in the region under the electrode and usually has a low capture threshold. On the contrary, anodal stimulation results in the accumulation of negatively charged ions at the myocardial–lead interface, which hyperpolarizes adjacent cells, so that the ability to trigger an action potential is paradoxical. However, anodal capture is routinely observed in cardiac pacing and usually requires a high pacing output, which can be explained by the bidomain cardiac tissue model that proposes to induce virtual cathodes at sites distant from the electrode to initiate depolarization.^17–19^ Therefore, the myocardium capture threshold depends on whether it is stimulated by the anode or cathode. Since the RVS capture threshold was higher than that of LVS and LBB in unipolar cathodal stimulation (Table 2), and the myocardial RVS capture by anodal stimulation in Tip Bipolar resulted in a further decrease in RVS myocardial capture threshold, the full fusion mode capture threshold was the highest. In Bilateral Cathodes configuration, all of the LBB\LVS and RVS were captured by cathodal stimulation, so the full fusion mode capture threshold was the lowest. In Ring Bipolar configuration, the LVS and LBB capture threshold increased as they were captured by anodal stimulation, but the full fusion mode capture threshold was still lower than that in Tip Bipolar configuration. The LVS and LBB capture threshold was lower than that of RVS in unipolar cathodal. In addition, the full fusion mode threshold was much more affected by the Tip Bipolar configuration than the Ring Bipolar configuration, likely because the tip electrodes had different surrounding ventricular myocardial areas with completely different morphology from those of the ring electrodes.

### Three new capture modes in Ring Bipolar and Bilateral Cathodes configurations

Only four capture modes were observed in conventional bipolar pacing mode (Tip Bipolar configuration), including full fusion mode (LVS+RVS+LBB), LVS+LBB, LBB, and LVS. Three new capture modes in Ring Bipolar and Bilateral Cathodes configurations were RVS+LBB, RVS+LVS, and RVS. It was speculated that LVS and LBB were captured by anodal stimulation in Ring Bipolar configuration, and the LVS and LBB capture threshold was increased and close to that of the RVS threshold. When the LVS capture threshold was higher than that of RVS and LBB, it changed to RVS+LBB capture mode. When the LBB capture threshold was higher than that of LVS and RVS, it changed to RVS+LVS capture mode. When the RVS capture threshold was lowest, it changed to a select RVS capture while further reducing the pacing output. In Bilateral cathodal configuration, the RVS, LVS, and LBB thresholds were also similar as they were all stimulated by the cathode. Thus, similar capture modes can be observed in Ring Bipolar configuration. On the contrary, RVS was captured by anodal stimulation in Tip Bipolar configuration, so the threshold was further increased and was much higher than that of LVS and LBB. This explained why only the LVS+LBB capture mode exists in Tip Bipolar configuration.

### P-QRS \ V6 RWPT \ V1 RWPT of full fusion mode

The present study compared the P-QRS, V6 RWPT, and V1 RWPT of seven capture modes. Full fusion mode had the shortest P-QRS, V6 RWPT, and V1 RWPT. The P-QRS duration of full fusion mode was compared to LB-QRS duration in patients with a native narrow or wide QRS. The results showed that the full fusion mode can achieve a short P-QRS in patients with a native wide QRS. Full fusion mode significantly improved interventricular dyssynchrony and RV depolarization durations compared to other capture modes in LBBP. Although, we also found there was a finding about bilateral bundle branch area pacing has been reported,^20^ which was similar to full fusion mode. But the LBB capture criteria was used in that study can not precisely confirm LBB capture as the direct evidence of LBB capture had not been reported at that time. The average V6 RWPT was about 82-84 ms in the study of bilateral bundle branch area pacing, which was longer than it was reported in our study (65-68 ms) and other study recently (64-69 ms) ^21^, it indicated there may had patients only capture LVS and without LBB capture. Full fusion mode was recognized by dynamic ECG and EGM maneuvers which can precisely confirm capture myocardial components in our study. So full fusion mode should be different from bilateral bundle branch area pacing.

### Clinical implications

The capture threshold is high in conventional bipolar pacing mode in order to achieve full fusion mode. The present study showed that changing the bilateral electrode pacing vector configuration can reduce the full fusion mode capture threshold, especially in Bilateral Cathode and Ring Bipolar configurations. The CRT device already has multiple electrode pacing vector configurations. If this function can be applied in a normal pacemaker device, more patients can benefit from full fusion mode as it can achieve a more physiological ventricular depolarization than conventional LBBP can.

## Limitations

Although it was demonstrated that changing the bilateral electrode pacing vector configuration can reduce the full fusion mode capture threshold, whether it can be used in a clinical setting remains unknown. The seven capture mode thresholds in three bilateral electrode pacing vector configurations were acute pacing parameters, long-term follow-up was necessary to confirm the threshold stable. Pacemaker manufacturers might need to make technical improvements to accommodate this technique. In addition, full fusion mode is a physiological pacing mode based on cardiac electrophysiological data. Whether it will result in a better hemodynamic response or less mechanical dyssynchrony in a clinical setting remains a question to be addressed. Clinical trials are necessary to further investigate the efficacy of this method. Changes in the LB lead EGM can help to distinguish RVS+LBB capture mode from full fusion mode since paced ECG morphology results were similar. There may be some difficulty in designating separate EGM in a few patients in Bilateral Cathodal mode as the EGM amplitude was low and may cause some deviation. Finally, the present study was performed at a single center and had a small sample size, which may cause bias in the statistical results.

## Conclusion

The present study showed that changing the bilateral electrode pacing vector configuration to Bilateral Cathodes and Ring Bipolar configurations can reduce the full fusion mode capture threshold compared to conventional bipolar pacing. There were seven capture modes in Ring Bipolar and Bilateral Cathodes during threshold testing, including full fusion mode (RVS+LVSP+LBB), semi fusion mode (LVS+LBB, RVS+LBB, and RVS+LVS), and select capture mode (LBB, LVS, and RVS). The full fusion mode had the shortest P-QRS, V6 RWPT, and V1 RWPT, which was a more physiological pacing mode than conventional LBBP.

## Data Availability

The data that support the findings of this study are available from the corresponding author, Longfu Jiang, MD, upon reasonable request.

---

**SUPPLEMENTARY FIGURE 1.**
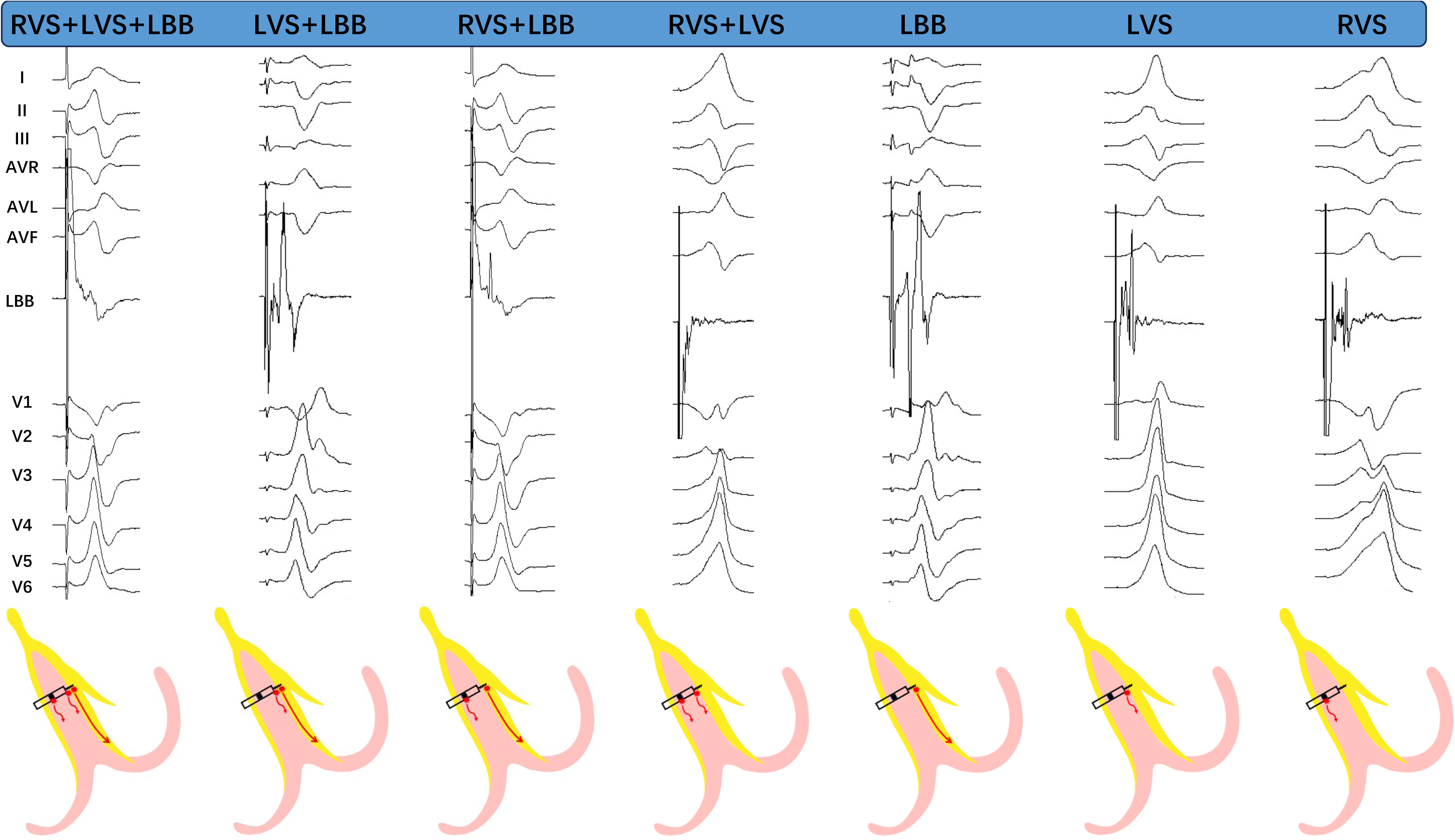
The changes in EGM and ECG during the threshold test of the three bilateral electrode pacing vector configurations.

